# Comparison of alcohol septal ablation and mavacamten in patients with obstructive hypertrophic cardiomyopathy: a propensity-matched large single-center study

**DOI:** 10.64898/2026.08.18.26360764

**Authors:** Jan Koelemen, Kay Becht, Christoph Reich, Ali Amr, Elham Kayvanpour, Steffen Rosskopf, Norbert Frey, Benjamin Meder, Farbod Sedaghat-Hamedani

**Author notes:** Corresponding author: PD Dr. med. Farbod Sedaghat-Hamedani, MD, FESC Department of Cardiology, Angiology and Pneumology Institute for Cardiomyopathies Heidelberg (ICH.) University of Heidelberg, Heidelberg University Hospital Im Neuenheimer Feld 410, 69120 Heidelberg Germany.

## Abstract

**Background:** Obstructive hypertrophic cardiomyopathy (oHCM) causes substantial symptom burden and impaired functional capacity. Mavacamten has emerged as a targeted pharmacologic treatment, whereas alcohol septal ablation (ASA) is an established septal reduction therapy (SRT). Direct comparative real-world data remain limited.

**Methods:** In this propensity-controlled observational study, longitudinal registry data from Heidelberg University Hospital were analyzed. Consecutive adults with oHCM, NYHA class ≥II symptoms, and a maximum LVOT gradient ≥50 mmHg treated with mavacamten or ASA were included. The cohort comprised 107 ASA- and 113 mavacamten-treated patients. Follow-up was performed at 6 and 12 months. The primary endpoint was a composite adverse clinical outcome including cardiovascular death, heart failure hospitalization, SRT, heart transplantation, ventricular assist device implantation, permanent pacemaker implantation for third-degree atrioventricular block, or decline in left ventricular ejection fraction to <40%.

**Results:** Both treatments showed significant improvement in NYHA class and LVOT gradient reduction over 12 months. Mean LVOT gradient decreased from 100.3 to 44.2 mmHg after ASA and from 85.7 to 18.4 mmHg with mavacamten at 12 months (both p<0.001). Between-group differences were not significant at 6 months, whereas residual LVOT gradient was lower with mavacamten at 12 months (p=0.004). NT-proBNP declined in both groups and was lower with mavacamten at both follow-up visits (both p<0.001). Third-degree atrioventricular block occurred more frequently after ASA (6.5% vs 0%, p=0.002). The composite endpoint occurred in 13 ASA-(12.1%) and 4 mavacamten-treated patients (3.5%) (p=0.003), with higher 1-year event-free survival in the mavacamten group (HR 0.19; 95%-CI 0.06-0.60; p=0.001).

**Conclusions:** In this real-world comparative study, both ASA and mavacamten improved symptoms and LVOT obstruction in oHCM. Mavacamten was associated with a more favorable short-term hemodynamic and safety profile at 12 months.

**Graphical abstract.:** Study design and principal findings of a propensity-matched single-center cohort study comparing alcohol septal ablation and mavacamten in patients with symptomatic obstructive hypertrophic cardiomyopathy.

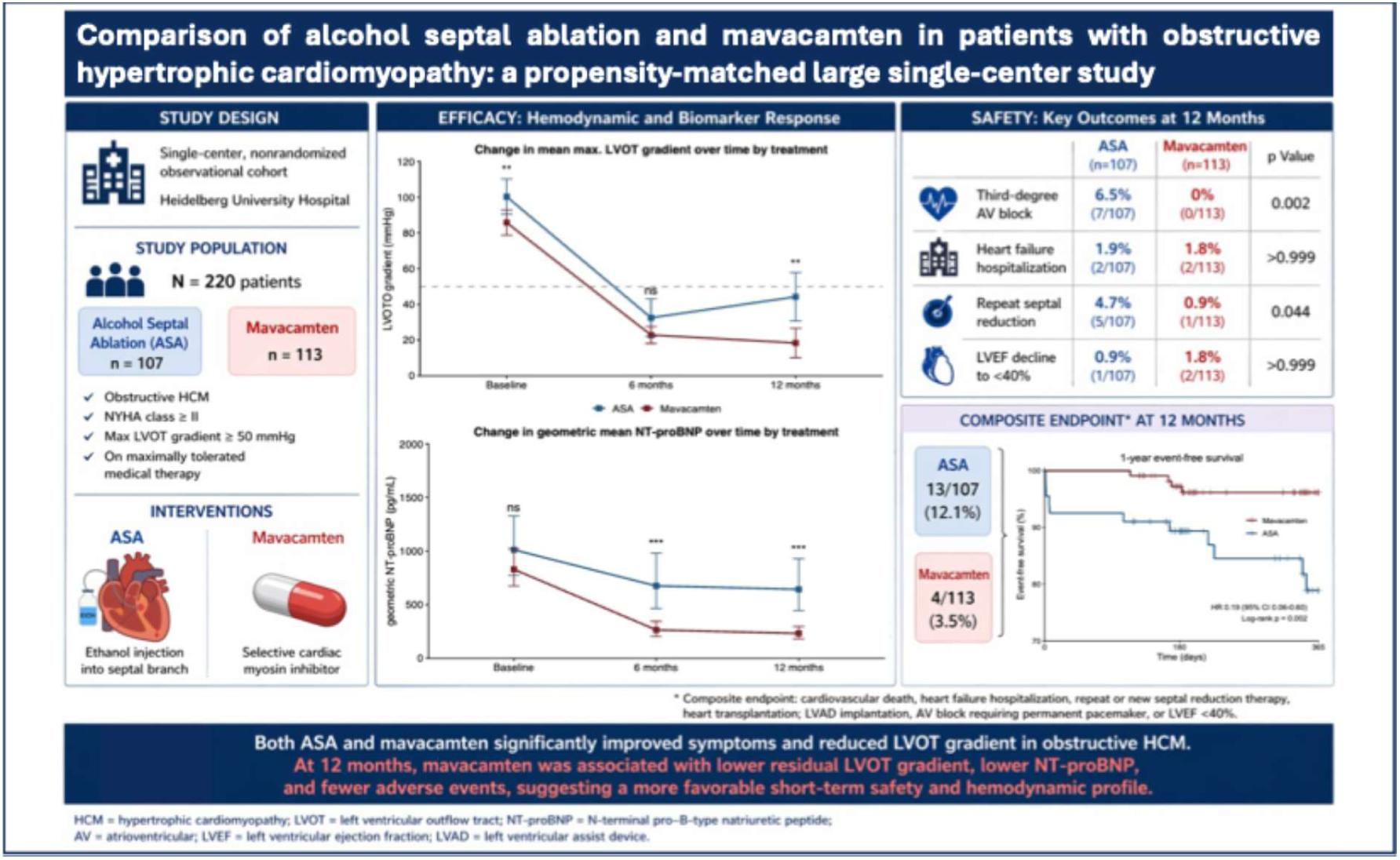

## Introduction

Hypertrophic cardiomyopathy (HCM) is the most common inherited myocardial disease, with a population prevalence of approximately 1:500. Left ventricular outflow tract obstruction (LVOTO) is a central determinant of symptoms and clinical progression in a substantial proportion of affected patients (1,2). The pathophysiology of this obstruction is multifactorial, rooted in sarcomeric mutations that lead to excessive actin-myosin cross-bridging and myocardial hypercontractility (3,4). This hyperdynamic state, combined with basal septal hypertrophy and systolic anterior motion (SAM) of the mitral valve apparatus, leads to LVOTO. Consequently, patients frequently suffer from exertional dyspnea, vertigo, chest pain, syncope, impaired quality of life and a heightened risk of heart failure progression. (1,2).

Contemporary management strategies focus on timely relief of obstruction, initially with negative inotropic therapy and, when symptoms persist, escalation to advanced pharmacologic treatment or septal reduction therapy (SRT) (1,2,5).

Surgical septal myectomy has served as the reference standard invasive treatment since the 1960s and remains highly effective when performed in expert centers (2,6,7). In the mid-1990s, catheter-based septal ablation emerged as a nonsurgical alternative, and subsequent refinement using myocardial contrast echocardiographic guidance improved target-vessel selection and procedural precision (8–10). Over the following two decades, alcohol septal ablation (ASA) matured from an innovative option to an established SRT for selected symptomatic patients with obstructive HCM, especially in Europe (1,2,11,12). Large observational series demonstrated durable hemodynamic and symptomatic improvement together with favorable long-term survival after ASA (11–33). At the same time, meta-analyses consistently suggest that ASA and myectomy achieve comparable long-term symptom relief, mortality and sudden death rates, with higher rates of pacemaker implantation after ASA (6,7,34–38).

More recently, advances in understanding the mechanisms of sarcomeric hypercontractility have opened the door to targeted pharmacotherapy. Mavacamten, a first-in-class cardiac myosin inhibitor (CMI), directly reduces actin-myosin cross-bridge formation and thereby addresses a proximal pathophysiologic driver of dynamic LVOTO rather than its anatomic consequence alone (1,2,39,40). In EXPLORER-HCM, mavacamten improved exercise capacity, New York Heart Association (NYHA) functional class, health status, and post-exercise left-ventricular outflow tract (LVOT) gradient in symptomatic obstructive HCM, establishing proof of concept for disease-specific pharmacotherapy (39). In VALOR-HCM, which enrolled patients already referred for SRT, mavacamten substantially reduced the proportion of patients who remained guideline-eligible for septal reduction or proceeded to invasive treatment after 16 weeks, and this benefit was sustained through 56 and 128 weeks (41–43). These data have already influenced clinical algorithms. The 2023 ESC guidelines position mavacamten as second-line therapy in adult patients with persistent resting or provoked LVOTO despite beta-blockers, calcium antagonists, and/or disopyramide, including as monotherapy when conventional agents are not tolerated (1). The 2024 AHA/ACC guidelines went further by recommending the addition of a myosin inhibitor, disopyramide, or SRT in persistently symptomatic obstructive HCM after first-line therapy with beta-blockers or non-dihydropyridine calcium channel blockers.(2). The double-blind, double-dummy randomized controlled trial (RCT) Maple-HCM recently questioned the use of beta-blockers and further supports the effectiveness of myosin-inhibition as the pivotal oHCM therapy (44).

Despite these major advances, the evidence base remains incomplete. Most invasive outcome data compare ASA with myectomy, whereas the CMI trials compared myosin inhibition with placebo rather than with established septal reduction strategies (6,7,39,43). Consequently, the practical decision between a fixed-duration or staged catheter intervention and a chronic, monitoring-intensive pharmacologic strategy is still guided more by extrapolation than by direct evidence. Indeed, only recently have small single-center retrospective analyses attempted a head-to-head comparison between ASA and mavacamten, underscoring both the clinical relevance of the question and the paucity of real-world comparative data (45). This gap is especially important in tertiary referral centers, where both therapies are increasingly available and where treatment selection must account for local expertise, baseline anatomy, patient comorbidity, follow-up infrastructure, and the acceptability of long-term drug surveillance.

To further address this gap in evidence, we conducted a single-center, non-randomized, observational and comparative cohort study using longitudinal registry data to evaluate 6- and 12-month efficacy and safety in symptomatic (New York Heart Association (NYHA) class ≥II) adults treated with either ASA or mavacamten.

## Methods

This single-center, nonrandomized, observational comparative cohort study was based on longitudinal registry data collected during routine clinical care at the Department of Cardiology, Institute for Cardiomyopathies (ICH), Heidelberg University Hospital, Heidelberg, Germany. The study was conducted in accordance with the principles of the Declaration of Helsinki and local ethical requirements. It is reported in accordance with the Strengthening the Reporting of Observational Studies in Epidemiology (STROBE) reporting guideline (46). The completed STROBE checklist is provided in the Supplementary Material.

Consecutive patients aged ≥18 years with primary hypertrophic cardiomyopathy, NYHA functional class II or higher, and a maximum LVOT gradient ≥50 mmHg were included if they underwent treatment with either ASA or mavacamten. Patients were excluded if left ventricular hypertrophy or LVOT obstruction was attributable to secondary causes. The study cohort included 107 patients treated with ASA before the commercial availability of mavacamten in Germany in August 2023 and 113 patients treated with mavacamten between August 2023 and August 2025. The ASA cohort was restricted to the pre-mavacamten era to avoid selection bias. After the introduction of mavacamten, the 2023 ESC Guidelines for the Management of Cardiomyopathies recommend septal reduction therapy predominantly for patients with persistent symptoms despite or after unsuccessful treatment with a cardiac myosin inhibitor (1). Therefore, inclusion of patients undergoing ASA after mavacamten became available would have enriched the ASA cohort with patients who had failed or were unsuitable for mavacamten therapy, thereby compromising comparability between the treatment groups. Baseline was defined as the last pre-treatment assessment obtained in routine care. No study-specific procedures were performed.

All data were derived from routine clinical care and the institutional registry. Baseline variables included demographic and clinical characteristics, medications, genotype status (when available), laboratory values, electrocardiographic findings, and echocardiographic parameters. Follow-up assessments were performed in the outpatient setting at 6 and 12 months after treatment initiation, with follow-up censored at 12 months for patients with longer observation periods.

The primary endpoint was a composite of major adverse cardiac events, defined as cardiovascular death, hospitalization for heart failure, repeat or new septal reduction therapy, heart transplantation, left ventricular assist device implantation, atrioventricular block requiring permanent pacemaker implantation, or a decline in left ventricular ejection fraction (LVEF) to <40%. Secondary endpoints included changes in NYHA functional class, LVOT gradient, NT-proBNP, high-sensitivity troponin T, left ventricular dimensions, LVEF, and electrocardiographic abnormalities.

Because treatment allocation was not randomized, a propensity score-matched analysis was performed as a sensitivity analysis and is presented in the supplementary material (Supplementary Table 1). Propensity scores were estimated using logistic regression with prespecified baseline variables (Supplementary Figure 1), followed by 1:1 nearest-neighbor matching without replacement with a caliper of 0.2 standard deviations of the logit of the propensity score. As both matched and unmatched analyses yielded similar results, the primary analyses were performed in the full and unmatched cohort.

Statistical analyses were performed using R version 4.5.2. Categorical variables are presented as counts and percentages and continuous variables as mean ±SD, mean with 95% confidence interval, or median with interquartile range, as appropriate. Between-group comparisons were performed using Welch’s t-test or the Wilcoxon rank-sum test; within-group longitudinal changes were assessed with the paired t-test or Wilcoxon signed-rank test, as appropriate. In the continuous-variable analyses, normality was assessed using the Shapiro-Wilk test. Categorical variables were compared with Fisher’s exact test. Time-to-event analyses were performed using Kaplan-Meier estimates and compared with the log-rank test, and Cox proportional-hazards models were used to estimate hazard ratios with 95% confidence intervals. All tests were two-sided, and p<0.05 was considered statistically significant.

## Results

### Study population and follow-up

The study included 220 patients, 107 treated with ASA and 113 with mavacamten. Overall follow-up was comparable between groups. Mean follow-up duration in the full cohort was 309.9±113.6 days, compared with 298.5±118.1 days after ASA and 316.6±110.9 days after mavacamten (p=0.369); the corresponding median follow-up times were 350, 344, and 354 days, respectively. At 12 months, 36% of patients in the mavacamten cohort had been up-titrated to a daily dose of ≥10 mg (Supplementary Figure 2). As expected in a registry-based analysis derived from routine care, completeness of follow-up varied across variables and time points and was generally lower in the historical ASA cohort.

### Baseline characteristics

Baseline characteristics are summarized in Table 1. The groups were broadly similar with respect to age, sex, body mass index, symptom burden, family history, genotype positivity among tested patients, prior syncope, non-sustained ventricular tachycardia (NSVT), baseline biomarker profile, left atrial size, systolic anterior motion, and background medical therapy. In both groups, most patients were in NYHA class II or III at study entry. At baseline, the concomitant use of beta-blockers, verapamil and loop diuretics did not differentiate significantly between the ASA and mavacamten groups (beta-blockers: 68% vs 77%, p=0.144; verapamil: 23% vs 17%, p=0.225; loop diuretics: 39% vs 27%, p=0.063).

**Table 1.**
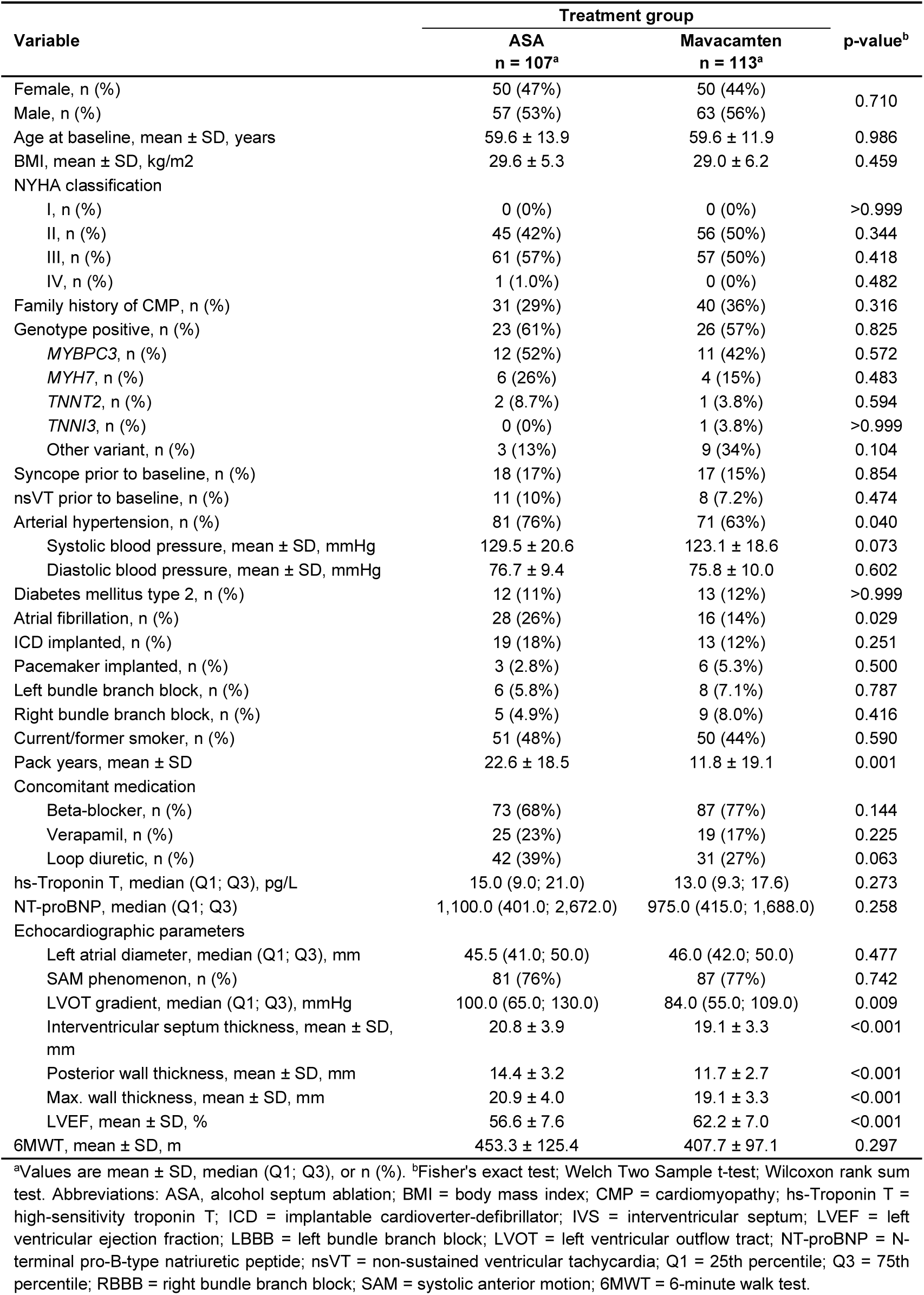
Unadjusted baseline characteristics of study cohort.

| Variable | Treatment group |  | p-value<sup>b</sup> |
| --- | --- | --- | --- |
|  | ASA<br>n = 107<sup>a</sup> | Mavacamten<br>n = 113<sup>a</sup> |  |
| Female, n (%) | 50 (47%) | 50 (44%) | 0.710 |
| Male, n (%) | 57 (53%) | 63 (56%) |  |
| Age at baseline, mean ± SD, years | 59.6 ± 13.9 | 59.6 ± 11.9 | 0.986 |
| BMI, mean ± SD, kg/m<sup>2</sup> | 29.6 ± 5.3 | 29.0 ± 6.2 | 0.459 |
| NYHA classification |  |  |  |
| I, n (%) | 0 (0%) | 0 (0%) | >0.999 |
| II, n (%) | 45 (42%) | 56 (50%) | 0.344 |
| III, n (%) | 61 (57%) | 57 (50%) | 0.418 |
| IV, n (%) | 1 (1.0%) | 0 (0%) | 0.482 |
| Family history of CMP, n (%) | 31 (29%) | 40 (36%) | 0.316 |
| Genotype positive, n (%) | 23 (61%) | 26 (57%) | 0.825 |
| MYBPC3, n (%) | 12 (52%) | 11 (42%) | 0.572 |
| MYH7, n (%) | 6 (26%) | 4 (15%) | 0.483 |
| TNNT2, n (%) | 2 (8.7%) | 1 (3.8%) | 0.594 |
| TNNI3, n (%) | 0 (0%) | 1 (3.8%) | >0.999 |
| Other variant, n (%) | 3 (13%) | 9 (34%) | 0.104 |
| Syncope prior to baseline, n (%) | 18 (17%) | 17 (15%) | 0.854 |
| nsVT prior to baseline, n (%) | 11 (10%) | 8 (7.2%) | 0.474 |
| Arterial hypertension, n (%) | 81 (76%) | 71 (63%) | 0.040 |
| Systolic blood pressure, mean ± SD, mmHg | 129.5 ± 20.6 | 123.1 ± 18.6 | 0.073 |
| Diastolic blood pressure, mean ± SD, mmHg | 76.7 ± 9.4 | 75.8 ± 10.0 | 0.602 |
| Diabetes mellitus type 2, n (%) | 12 (11%) | 13 (12%) | >0.999 |
| Atrial fibrillation, n (%) | 28 (26%) | 16 (14%) | 0.029 |
| ICD implanted, n (%) | 19 (18%) | 13 (12%) | 0.251 |
| Pacemaker implanted, n (%) | 3 (2.8%) | 6 (5.3%) | 0.500 |
| Left bundle branch block, n (%) | 6 (5.8%) | 8 (7.1%) | 0.787 |
| Right bundle branch block, n (%) | 5 (4.9%) | 9 (8.0%) | 0.416 |
| Current/former smoker, n (%) | 51 (48%) | 50 (44%) | 0.590 |
| Pack years, mean ± SD | 22.6 ± 18.5 | 11.8 ± 19.1 | 0.001 |
| Concomitant medication |  |  |  |
| Beta-blocker, n (%) | 73 (68%) | 87 (77%) | 0.144 |
| Verapamil, n (%) | 25 (23%) | 19 (17%) | 0.225 |
| Loop diuretic, n (%) | 42 (39%) | 31 (27%) | 0.063 |
| hs-Troponin T, median (Q1; Q3), pg/L | 15.0 (9.0; 21.0) | 13.0 (9.3; 17.6) | 0.273 |
| NT-proBNP, median (Q1; Q3) | 1,100.0 (401.0; 2,672.0) | 975.0 (415.0; 1,688.0) | 0.258 |
| Echocardiographic parameters |  |  |  |
| Left atrial diameter, median (Q1; Q3), mm | 45.5 (41.0; 50.0) | 46.0 (42.0; 50.0) | 0.477 |
| SAM phenomenon, n (%) | 81 (76%) | 87 (77%) | 0.742 |
| LVOT gradient, median (Q1; Q3), mmHg | 100.0 (65.0; 130.0) | 84.0 (55.0; 109.0) | 0.009 |
| Interventricular septum thickness, mean ± SD, mm | 20.8 ± 3.9 | 19.1 ± 3.3 | <0.001 |
| Posterior wall thickness, mean ± SD, mm | 14.4 ± 3.2 | 11.7 ± 2.7 | <0.001 |
| Max. wall thickness, mean ± SD, mm | 20.9 ± 4.0 | 19.1 ± 3.3 | <0.001 |
| LVEF, mean ± SD, % | 56.6 ± 7.6 | 62.2 ± 7.0 | <0.001 |
| 6MWT, mean ± SD, m | 453.3 ± 125.4 | 407.7 ± 97.1 | 0.297 |

<sup>a</sup>Values are mean ± SD, median (Q1; Q3), or n (%). <sup>b</sup>Fisher's exact test; Welch Two Sample t-test; Wilcoxon rank sum test. Abbreviations: ASA, alcohol septum ablation; BMI = body mass index; CMP = cardiomyopathy; hs-Troponin T = high-sensitivity troponin T; ICD = implantable cardioverter-defibrillator; IVS = interventricular septum; LVEF = left ventricular ejection fraction; LBBB = left bundle branch block; LVOT = left ventricular outflow tract; NT-proBNP = N-terminal pro-B-type natriuretic peptide; nsVT = non-sustained ventricular tachycardia; Q1 = 25th percentile; Q3 = 75th percentile; RBBB = right bundle branch block; SAM = systolic anterior motion; 6MWT = 6-minute walk test.

Several baseline differences were observed. Patients undergoing ASA more frequently had hypertension (76% vs 63%, p=0.040) and atrial fibrillation (26% vs 14%, p=0.029), and had a greater smoking burden (22.6 ± 18.5 vs 11.8 ± 19.1 pack-years, p<0.001). The ASA cohort also showed a more pronounced obstructive and structural phenotype, with a higher median LVOT gradient (100 [65-130] mmHg vs 84 [55-109] mmHg, p=0.009), greater mean interventricular septal thickness (20.8 ± 3.9 mm vs 19.1 ± 3.3 mm, p<0.001), greater posterior wall thickness (14.4 ± 3.2 mm vs 11.7 ± 2.7 mm, p<0.001), greater mean maximal wall thickness (20.9 ± 4.0 mm vs 19.1 ± 3.3 mm, p<0.001), and a slightly lower, yet normal mean LVEF (56.6 ± 7.6% vs 62.2 ± 7.0%, p<0.001).

### Functional status

Both treatment strategies were associated with a clear improvement in functional status over time (Figure 1 & 2, Supplementary Figure 3). The alluvial plots for patient trajectories demonstrated a shift toward lower NYHA functional classes at 6 and 12 months in both groups (Supplementary Figure 3). Direct between-group comparisons of the degree of improvement or worsening showed no significant differences at either follow-up time point (Figure 2; 6 months: p = 0.480; 12 months: p = 0.932).

**Figure 1.**
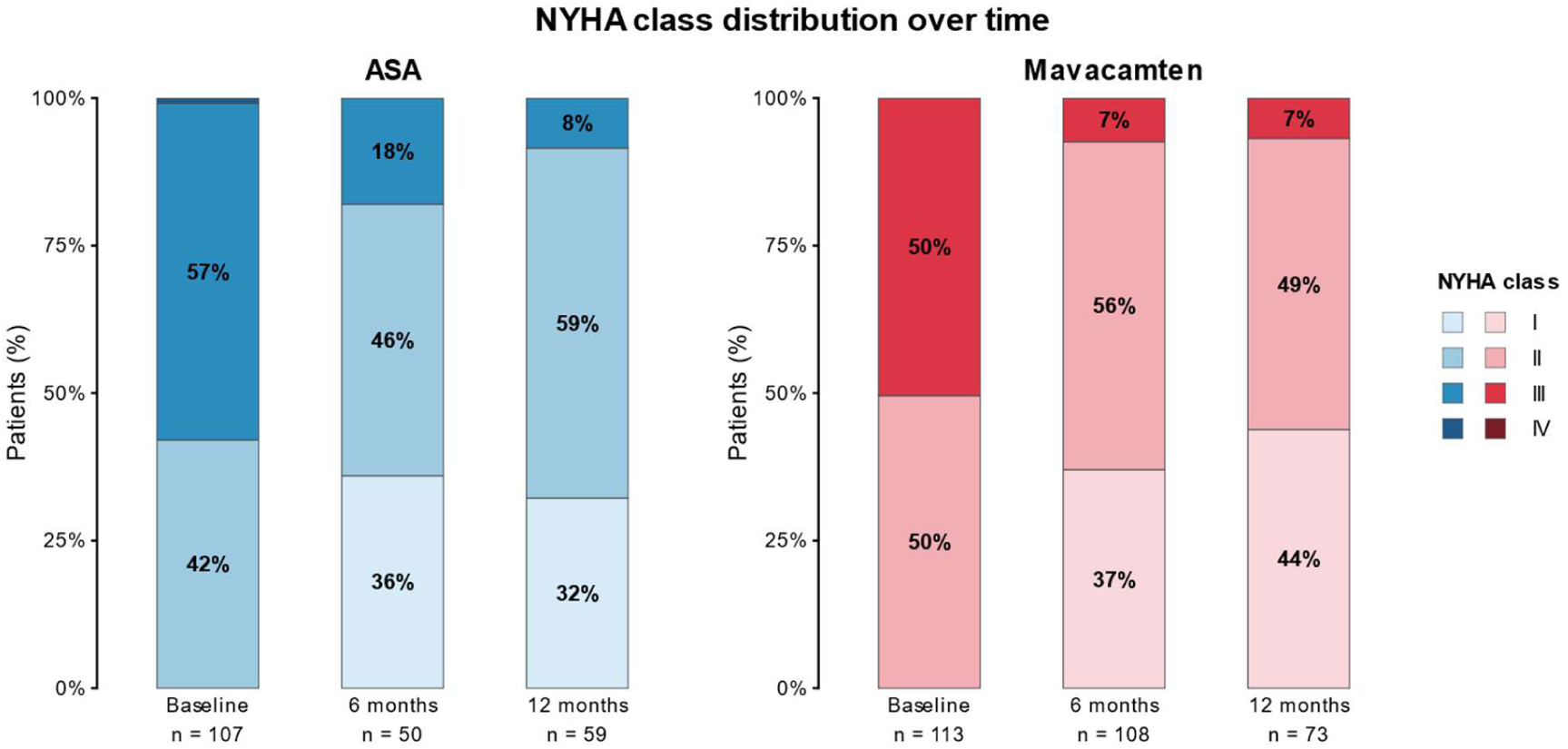
New York Heart Association (NYHA) functional class distribution at baseline, 6 months, and 12 months after alcohol septal ablation (ASA, left) and mavacamten therapy (right). Stacked bar charts show the proportion of patients in each NYHA class at each timepoint. All available observations were included for each follow-up assessment, resulting in varying sample sizes across timepoints. Patient numbers are shown below each bar.

**Figure 2.**
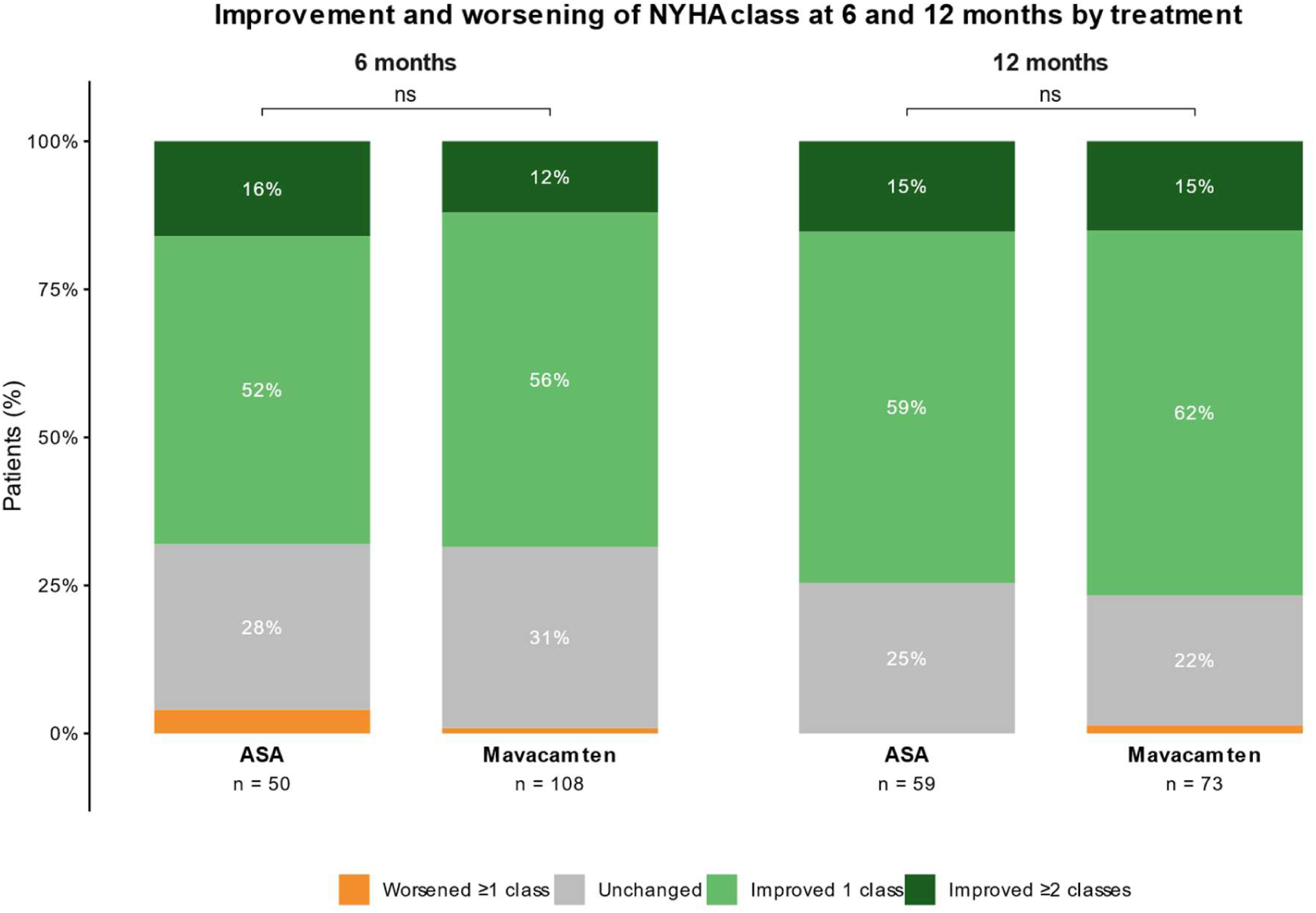
Improvement and worsening of NYHA functional class at 6 and 12 months according to treatment group. Stacked bar charts show the proportion of patients with worsening (≥1 class), unchanged status, improvement by 1 class, and improvement by ≥2 classes following ASA and mavacamten therapy. Between-group comparisons of the ordinal distribution of NYHA functional class changes were performed using Fisher’s exact test. No significant differences between treatment groups were observed at either timepoint (6 months: p = 0.480; 12 months: p = 0.932).

### LVOT gradient and biomarker response

Both treatments were associated with marked reductions in LVOT gradient (Figure 3A-B). After ASA, mean LVOT gradient decreased from 100.3 mmHg (95% CI, 90.4-110.2) at baseline to 32.5 mmHg (95% CI, 22.0-43.1) at 6 months and 44.2 mmHg (95% CI, 30.7-57.7) at 12 months (both p<0.001 vs baseline). After mavacamten, mean LVOT gradient decreased from 85.7 mmHg (95% CI, 78.7-92.8) to 22.8 mmHg (95% CI, 18.1-27.6) at 6 months and 18.4 mmHg (95% CI, 10.2-26.6) at 12 months (both p<0.001 vs baseline). Between-group differences were no longer evident at 6 months (p=0.472), whereas at 12 months the LVOT gradient remained lower in the mavacamten group (p=0.004). Within the mavacamten cohort, an additional reduction was observed between 6 and 12 months (p=0.017), whereas no further significant change occurred in the ASA cohort (p=0.567).

**Figure 3.**
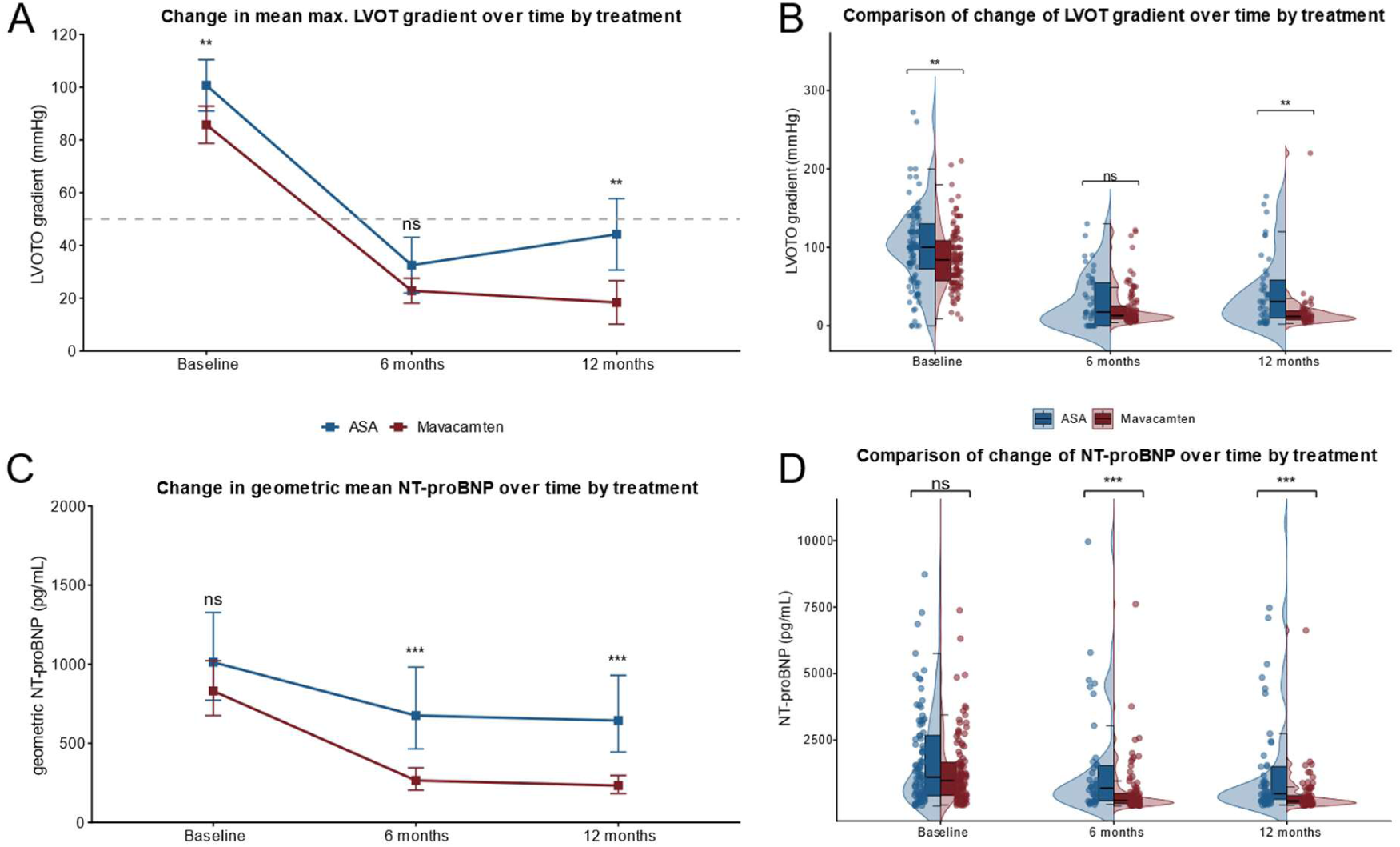
Changes in maximal left ventricular outflow tract (LVOT) gradient and N-terminal pro-B-type natriuretic peptide (NT-proBNP) over time according to treatment. **(A)** Mean LVOT gradient (±95% confidence interval) at baseline, 6 months, and 12 months in patients treated with alcohol septal ablation (ASA) and mavacamten. Patient numbers (ASA / mavacamten) were n=107 / 113 at baseline, n=42 / 101 at 6 months, and n=42 / 51 at 12 months. Both groups showed a marked reduction from baseline, with sustained improvement over time. The dashed horizontal line indicates the clinically relevant threshold of 50 mmHg. **(B)** Distribution of LVOT gradients at each time point shown as raincloud plots (half-violin, boxplot, and individual data points). Patient numbers (ASA / mavacamten) were n=107 / 113 at baseline, n=42 / 101 at 6 months, and n=42 / 51 at 12 months. Between-group comparisons were performed using Wilcoxon rank-sum tests. **(C)** Geometric mean NT-proBNP values (±95% confidence intervals) at baseline, 6 months, and 12 months in patients treated with ASA and mavacamten. Only positive values were included for calculation of the geometric mean. Patient numbers (ASA / mavacamten) were n=97 / 111 at baseline, n=42 / 96 at 6 months, and n=53 / 73 at 12 months. Both groups showed a reduction over time, with a more pronounced early decline in the mavacamten group. **(D)** Distribution of NT-proBNP values at each time point shown as raincloud plots (half-violin, boxplot, and individual data points). Patient numbers (ASA / mavacamten) were n=97 / 111 at baseline, n=42 / 96 at 6 months, and n = 53 / 73 at 12 months. Values exceeding 10,000 pg/mL were excluded from visualization (baseline: ASA n=3/97 [3.1%], mavacamten n=1/111 [0.9%]; 6 months: ASA n=0/42 [0%], mavacamten n=2/96 [2.1%]; 12 months: ASA n=2/53 [3.8%], mavacamten n=0/73 [0%]). ns, not significant; *p<0.05; **p<0.01; ***p<0.001.

NT-proBNP declined significantly in both groups (Figure 3 C-D). In the ASA cohort, NT-proBNP decreased from 1013 ng/L (95% CI, 773-1372) at baseline to 675 ng/L (95% CI, 465-981) at 6 months and 643 ng/L (95% CI, 445-930) at 12 months, with significant reductions from baseline (p=0.008 and p<0.001, respectively), but no further change between 6 and 12 months (p=0.556). In the mavacamten cohort, NT-proBNP decreased from 831 ng/L (95% CI, 675-1022) at baseline to 265 ng/L (95% CI, 204-345) at 6 months and 233 ng/L (95% CI, 182-296) at 12 months (both p<0.001 vs baseline), with a further reduction between 6 and 12 months (p=0.006). Baseline NT-proBNP values did not differ significantly between groups (p=0.258), whereas NT-proBNP was lower with mavacamten at both follow-up visits (both p<0.001).

### Structural and electrocardiographic changes

Both therapies were associated with reductions in septal and posterior wall thickness over time (Supplementary Figure 4A & 4B). Maximal wall thickness declined significantly from baseline in both groups at 6 and 12 months as well (all p<0.001). The baseline between-group difference was no longer significant at 6 months (p=0.192), but a modest difference was again present at 12 months, with lower values in the mavacamten group: 18.1 [16.8-19.3] mm after ASA vs 16.2 [15.5-16.8] mm after mavacamten; p=0.009. A similar pattern was seen for interventricular septal thickness (Supplementary Figure 4A). Although septal thickness was greater in the ASA group at baseline, the between-group difference was not significant at 6 months (p=0.236). At 12 months, septal thickness remained slightly lower with mavacamten. Posterior wall thickness was lower in the mavacamten group at baseline and remained lower at both follow-up visits (all p<0.001), although both groups showed significant reductions from baseline (Supplementary Figure 4B).

Electrocardiographic voltage, assessed by the Sokolow-Lyon index, decreased significantly in both groups at 6 and 12 months compared with baseline (all p<0.001), without significant between-group differences (Supplementary Figure 4C). By contrast, exercise-based measures showed no consistent changes. No significant changes in six-minute walk distance or ergometry parameters were observed over time, however data were available for only a limited subset of patients (Supplementary Figure 5).

LVEF was higher at baseline in the mavacamten group than in the ASA group (mean 62.2% vs 56.6%, p<0.001), but declined significantly in the mavacamten group at 6 and 12 months (57.7% and 57.4%, respectively; both p<0.001 vs baseline), with a small additional difference also observed between 6 and 12 months (p=0.020), whereas LVEF remained stable in the ASA group over time (56.6%, 60.8%, and 59.7% at baseline, 6 months, and 12 months, respectively; all p>0.20) (Supplementary Figure 4D). Accordingly, between-group differences were no longer significant at follow-up (6 months: p=0.053; 12 months: p=0.784).

### Clinical events

Clinical events are summarized in Table 2. Third-degree atrioventricular block occurred more frequently after ASA than after mavacamten (6.5% vs 0%, p=0.002). Heart failure hospitalization was infrequent and did not differ significantly between groups (1.9% vs 1.8%, p>0.999). A reduction in LVEF to <40% was uncommon in both groups (0.9% after ASA and 1.8% after mavacamten, p>0.999). No sudden cardiac deaths, cardiovascular deaths, heart transplantation or left ventricular assist device (LVAD) implantations occurred during follow-up in either group. New or repeat septal reduction procedures were more frequent after ASA (4.7% vs 0.9%, p=0.044). New myectomy was rare in both groups (both 0.9%, p>0.999). Temporary discontinuation of mavacamten occurred in 3 patients (2.8%).

**Table 2.** Adverse events during follow-up.

| Variable | Treatment group |  | p-value |
| --- | --- | --- | --- |
|  | ASA<br>n = 107 | Mavacamten<br>n = 113 |  |
| Follow-up time, mean ± SD, days | 299 ± 118 | 317 ± 111 | 0.369 |
| 3rd degree AV-Block, n (%) | 7 (6.5%) | 0 (0%) | 0.002 |
| New ASA after initial treatment, n (%) | 5 (4.7%) | 1 (0.9%) | 0.044 |
| New myectomy after initial treatment, n (%) | 1 (0.9%) | 1 (0.9%) | >0.999 |
| Temporary mavacamten discontinuation, n (%) | NA | 3 (2.8%) | >0.999 |
| Heart failure hospitalization, n (%) | 2 (1.9%) | 2 (1.8%) | >0.999 |
| Sudden cardiac death, n (%) | 0 (0%) | 0 (0%) | >0.999 |
| CV death, n (%) | 0 (0%) | 0 (0%) | >0.999 |
| Heart transplantation or LVAD, n (%) | 0 (0%) | 0 (0%) | >0.999 |
| LVEF reduction to <40%, n (%) | 1 (0.9%) | 2 (1.8%) | >0.999 |
| Composite adverse endpoint, n (%) | 13 (12.1%) | 4 (3.5%) | 0.003 |

Data are shown as n (%), with percentages based on complete-case data. Comparisons were performed using Fisher's exact test. For the composite endpoint, each patient contributed only once regardless of multiple events, and no weighting was applied to individual components. Abbreviations: ASA, alcohol septum ablation; AV, atrioventricular; CV, cardiovascular; HTX, heart transplantation; LVAD, left ventricular assist device; LVEF, left ventricular ejection fraction.

Overall, the predefined composite endpoint occurred in 13 patients (12.1%) in the ASA group and in 4 patients (3.5%) in the mavacamten group (p=0.003). Kaplan-Meier analysis showed significantly better event-free survival with mavacamten over 1 year, with a hazard ratio of 0.19 (95% CI, 0.06 to 0.60; log-rank p=0.002) (Figure 4).

**Figure 4.**
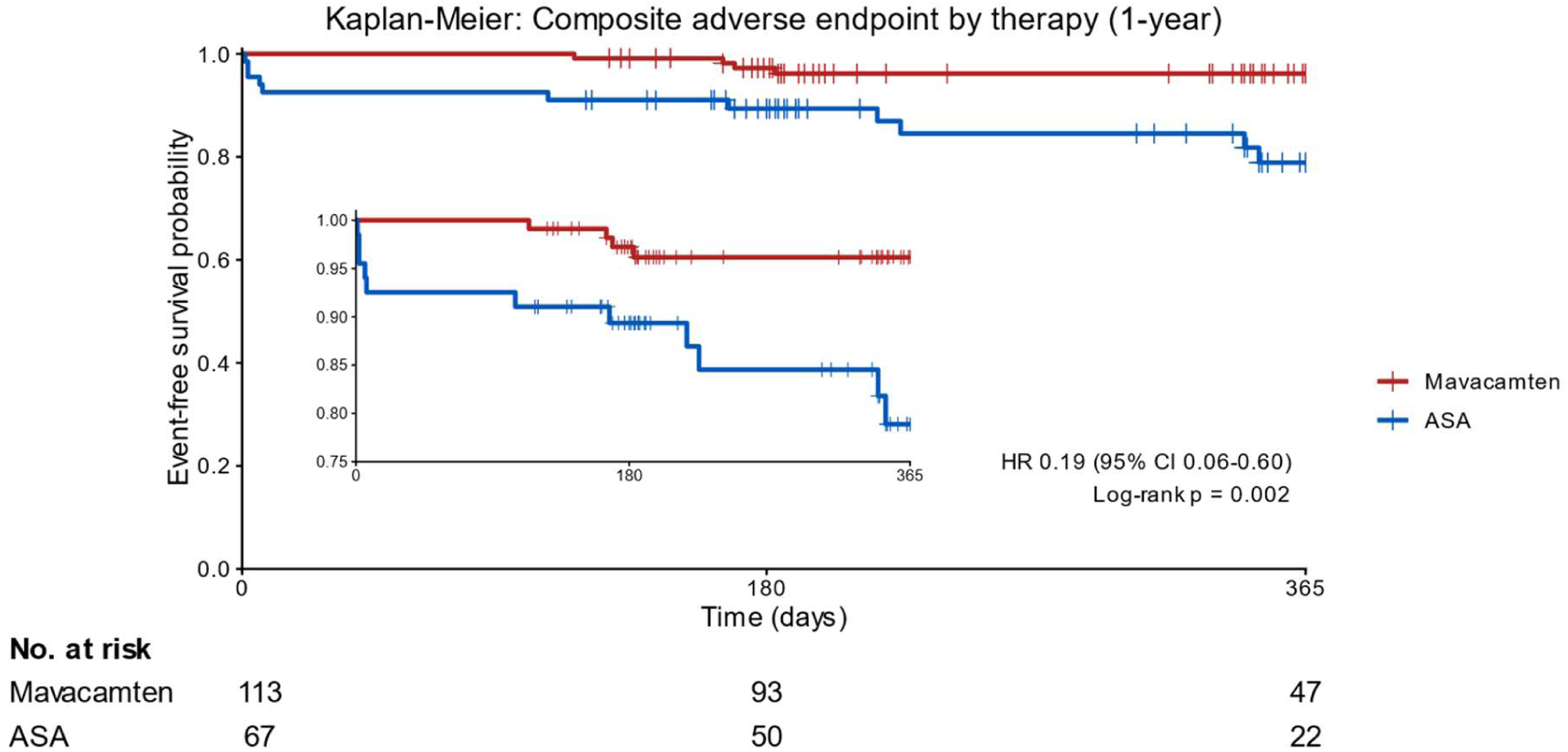
Kaplan-Meier estimates of event-free survival for the predefined composite clinical endpoint over 1 year according to treatment group. The composite endpoint comprised heart failure hospitalization, sudden cardiac death, cardiovascular death, heart transplantation or left ventricular assist device implantation, LVEF reduction <40%, new myectomy or ASA, and 3rd-degree atrioventricular block. Patients were counted once for the composite endpoint irrespective of multiple events. Of the 107 ASA patients, 67 had complete time-to-event information and were therefore included in the survival analysis. Mavacamten was associated with a significantly lower risk of the composite adverse endpoint compared with ASA (hazard ratio 0.19, 95% CI 0.06-0.60; log-rank p=0.002). Tick marks indicate censored observations; numbers at risk are shown below the x-axis.

## Discussion

In this single-center observational comparison of ASA and mavacamten in symptomatic obstructive hypertrophic cardiomyopathy, both treatment strategies were associated with meaningful clinical improvement over 12 months. Functional status improved in both groups, and both therapies produced substantial reductions in LVOT obstruction. At the same time, the pattern of response was not identical. The hemodynamic effect of ASA was marked early after treatment, whereas mavacamten was associated with a lower residual LVOT gradient and lower NT-proBNP at 12 months. In addition, the lower rate of the predefined composite endpoint with mavacamten was driven primarily by treatment-related events, particularly high-grade atrioventricular block requiring permanent pacing after ASA, rather than by differences in mortality or advanced heart failure events.

These findings should be interpreted within the contemporary treatment framework for obstructive hypertrophic cardiomyopathy. Both the 2023 ESC cardiomyopathy guidelines and the 2024 AHA/ACC hypertrophic cardiomyopathy guideline position septal reduction therapy as an established option for patients with relevant LVOT obstruction and persistent symptoms despite medical therapy, while also incorporating prior cardiac myosin inhibition into the therapeutic algorithm for selected symptomatic patients (1,2). This approach is also supported by a recently published ESC expert consensus paper (5). The present data support this evolving model of care rather than a binary view of either treatment as universally preferable.

The most direct comparison in the current literature is the recent study by Samhan et al., which examined ASA and mavacamten in a single-center retrospective cohort and found that both treatments were associated with substantial reductions in LVOT gradient and improvement in NYHA class by 32 weeks (45). A particularly relevant observation from that study was the different temporal profile of benefit: ASA achieved its maximal gradient reduction by 16 weeks, whereas mavacamten reached its peak effect later, at 32 weeks (45). The present findings are highly consistent with that pattern. ASA produced a pronounced early hemodynamic response, whereas mavacamten showed continued improvement between 6 and 12 months, with a further decline in both LVOT gradient and NT-proBNP. This temporal difference is clinically relevant, because short-term comparisons may underestimate the full effect of myosin inhibition when dose titration and longitudinal surveillance are part of routine care. Besides the higher patient numbers, the current study also investigated the dynamics of structural and electrocardiographic parameters and found signs of reverse remodeling in both therapies -which was often debated to be the case after ASA. At the same time, the present study extends the currently available evidence from comparison studies in several other important respects (45). First, the cohort was substantially larger and numerically balanced. Follow-up extended to 12 months rather than 32 weeks. The analysis was not limited to hemodynamic and symptomatic parameters, but also incorporated biomarkers, structural echocardiographic remodeling, electrocardiographic changes, and a prespecified clinical composite endpoint. Genetic information, serving as the etiologic basis for HCM classification, was available in a relevant proportion of patients, was well balanced between treatment groups, and was broadly consistent with the genetic yield reported in contemporary HCM literature (1,2,47). The present study therefore addresses a broader question than whether both treatments reduce obstruction; it evaluates how they compare across functional, biological, structural, and event-based domains in real-world practice.

The biomarker findings deserve particular attention. NT-proBNP declined in both groups but was lower with mavacamten at both follow-up visits, with a further reduction between 6 and 12 months only in the mavacamten cohort. This pattern is consistent with the broader mavacamten literature. In VALOR-HCM, mavacamten reduced the proportion of patients proceeding to or remaining eligible for septal reduction therapy after 16 weeks (43), and these effects remained durable through longer follow-up (41,42). Long-term extension data have likewise shown sustained improvements in obstruction, symptoms, and biomarker profiles over time (48,49). Although the present study was not designed to replicate a trial population, the continued fall in LVOT gradient and NT-proBNP at 12 months is directionally concordant with those observations and suggests that the physiological effect of mavacamten may continue to evolve beyond the early treatment phase.

The event analysis warrants a more careful interpretation than a straightforward claim of superiority. In this cohort, the lower 1-year composite event rate with mavacamten appeared to be driven primarily by treatment-related procedural events and repeat interventions, most notably third-degree atrioventricular block after ASA. That finding is biologically plausible and consistent with the established ASA literature. Large observational series have shown that alcohol septal ablation is associated with excellent long-term survival, but at the cost of a meaningful pacemaker burden and a non-negligible reintervention rate (11,12). In the Euro-ASA registry analysis by Veselka et al., 9.4% of patients required permanent pacemaker implantation within 30 days after ASA, comparable to the event rate observed in this cohort [9]. Meta-analytic comparisons of septal myectomy and ASA have reached similar conclusions: mortality is broadly comparable, whereas pacemaker implantation and repeat septal reduction procedures are more frequent after ASA [10,11]. Seen in that context, the lower composite event rate with mavacamten in the present analysis is clinically relevant, but it should not be overinterpreted as evidence of superior protection from hard long-term cardiovascular outcomes. To address potential selection bias, it may be noted that approximately 95% of patients in the ASA cohort underwent treatment before the market introduction of mavacamten, making physician-driven preference for one of the two strategies unlikely to have substantially influenced treatment allocation in most cases.

Several aspects strengthen the present study. Propensity-score matching was performed as a sensitivity analysis, but baseline characteristics in the overall cohort were already reasonably balanced, and the matched analyses did not materially differ from those of the full population (Supplementary Tables 1-3, Supplementary Figures 1, 6). For that reason, the unmatched cohort was used for the primary analysis to preserve sample size and clinical interpretability. The consistency between the matched and unmatched analyses supports the robustness of the principal findings and reflects real-world circumstances. It addresses a clinically timely question for which direct comparative real-world data remain limited. The cohort was comparatively large for a single-center study, with equal treatment group sizes and follow-up assessments at 6 and 12 months. In contrast to prior work (45), the present analysis moved beyond LVOT gradient and NYHA class by incorporating biomarkers, structural remodeling, electrocardiographic changes, and a clinically meaningful composite endpoint. In addition, all data were derived from routine care, which enhances the real-world relevance of the findings. Finally, the concordance between the full-cohort and propensity-matched analyses makes it less likely that the overall conclusions were driven by measurable baseline imbalance alone.

## Conclusion

Results from this study suggest that both ASA and mavacamten provide meaningful clinical benefit in obstructive hypertrophic cardiomyopathy. Mavacamten may offer a more favorable short-term safety profile and a more sustained reduction in LVOT gradient and NT-proBNP over 12 months compared to ASA. Whether these differences translate into superior long-term clinical outcomes will require longer follow-up and, ideally, prospective and randomized comparative data.

Both treatment strategies will continue to have an important place in the management of obstructive HCM. Preserving expert ASA programs at specialized centers remains crucial to ensure access to a highly effective interventional alternative to medical therapy, which may soon be expanded by next-generation myosin inhibitors such as Aficamten (50). Importantly, ASA will likely remain indispensable for nonresponders and for patients in whom myosin inhibition is not appropriate, including pregnant women, women with a desire for pregnancy, and patients with impaired left ventricular systolic function.

## Limitations

This study has several limitations. The study was observational and nonrandomized, and residual confounding cannot be excluded despite the consistency between the full-cohort and propensity-matched analyses. The two treatment groups arose from different treatment eras, with ASA performed over a longer historical interval and mavacamten used in a more contemporary setting. Differences in referral patterns, imaging practice, monitoring intensity, and adjunctive management may therefore have influenced the results.

Follow-up was censored after 12 months, which is sufficient to compare early symptom relief, gradient reduction, biomarker response, and treatment-related adverse events, but not long-term durability, late reintervention, or survival.

The composite endpoint included treatment-specific components such as pacemaker implantation and repeat septal reduction therapy. These are clinically important outcomes, but they also favor a noninvasive strategy by design and should therefore be interpreted differently from death or advanced heart failure events. As in any registry-based analysis from routine care, completeness of follow-up varied across variables and time points, particularly in the historical cohort. Finally, this was a single-center study, and external generalizability may be limited.

## Data Availability

The data underlying this study are not publicly available.

## Abbreviations

ACC: American College of Cardiology;
AHA: American Heart Association;
ASA: alcohol septal ablation;
CI: confidence interval;
ESC: European Society of Cardiology;
HCM: hypertrophic cardiomyopathy;
ICH: Institute for Cardiomyopathies;
LV: left ventricular;
LVAD: left ventricular assist device;
LVEF: left ventricular ejection fraction;
LVOT: left ventricular outflow tract;
LVOTO: left ventricular outflow tract obstruction;
MR: mitral regurgitation;
NT-proBNP: N-terminal pro-B-type natriuretic peptide;
NYHA: New York Heart Association;
SAM: systolic anterior motion;
SRT: septal reduction therapy;
TASH: transcoronary ablation of septal hypertrophy.

Trial names used in the manuscript include EXPLORER-HCM and VALOR-HCM.

## Acknowledgements

CR is funded by the Clinician Scientist Program of Heidelberg University, Faculty of Medicine. Artificial intelligence (ChatGPT-5.4, OpenAI) was used to assist in improving the readability of the manuscript, assisting in the composition of the central illustration, and supporting coding tasks in R. All content was reviewed, validated, and approved by the authors, who take full responsibility for the accuracy and integrity of the work.

## References

1. Arbelo E, Protonotarios A, Gimeno JR et al. 2023 ESC Guidelines for the management of cardiomyopathies: Developed by the task force on the management of cardiomyopathies of the European Society of Cardiology (ESC). European Heart Journal 2023;44:3503–3626.

2. Ommen SR, Ho CY, Asif IM et al. 2024 AHA/ACC/AMSSM/HRS/PACES/SCMR Guideline for the Management of Hypertrophic Cardiomyopathy: A Report of the American Heart Association/American College of Cardiology Joint Committee on Clinical Practice Guidelines. Circulation 2024;149:e1239–e1311.

3. Amr A, Kayvanpour E, Reich C et al. Assessing the Applicability of Cardiac Myosin Inhibitors for Hypertrophic Cardiomyopathy Management in a Large Single Center Cohort. Rev Cardiovasc Med 2024;25:225.

4. Sedaghat-Hamedani F, Kayvanpour E, Meder B. Targeting the Sarcomere: Myosin Inhibitors as the Revolutionary Game Changer in Hypertrophic Cardiomyopathy. Rev Cardiovasc Med 2026;27:47341.

5. Meder B, Coats CJ, Leinwand LA et al. EJHF expert consensus statement on the diagnosis and management of hypertrophic cardiomyopathy. Eur J Heart Fail 2026;28:299–314.

6. Bytyci I, Nistri S, Morner S, Henein MY. Alcohol Septal Ablation versus Septal Myectomy Treatment of Obstructive Hypertrophic Cardiomyopathy: A Systematic Review and Meta-Analysis. J Clin Med 2020;9:3062.

7. Osman M, Kheiri B, Osman K et al. Alcohol septal ablation vs myectomy for symptomatic hypertrophic obstructive cardiomyopathy: Systematic review and meta-analysis. Clin Cardiol 2019;42:190–197.

8. Sigwart U. Non-surgical myocardial reduction for hypertrophic obstructive cardiomyopathy. Lancet 1995;346:211–4.

9. Faber L, Meissner A, Ziemssen P, Seggewiss H. Percutaneous transluminal septal myocardial ablation for hypertrophic obstructive cardiomyopathy: long term follow up of the first series of 25 patients. Heart 2000;83:326–31.

10. Seggewiss H, Faber L, Gleichmann U. Percutaneous transluminal septal ablation in hypertrophic obstructive cardiomyopathy. Thorac Cardiovasc Surg 1999;47:94–100.

11. Batzner A, Pfeiffer B, Neugebauer A, Aicha D, Blank C, Seggewiss H. Survival After Alcohol Septal Ablation in Patients With Hypertrophic Obstructive Cardiomyopathy. J Am Coll Cardiol 2018;72:3087–3094.

12. Veselka J, Liebregts M, Cooper R et al. Outcomes of Patients With Hypertrophic Obstructive Cardiomyopathy and Pacemaker Implanted After Alcohol Septal Ablation. JACC Cardiovasc Interv 2022;15:1910–1917.

13. Li P, Xue Y, Sun J et al. Outcome of alcohol septal ablation in mildly symptomatic patients with hypertrophic obstructive cardiomyopathy: A comparison with medical therapy. Clin Cardiol 2021;44:1409–1415.

14. Bassim E-S, Rick AN, Mayra G, Charanjit SR, Mackram FE. Alcohol septal ablation in patients with concomitant hypertrophic cardiomyopathy and aortic valvular stenosis. Catheter Cardiovasc Interv 2020;95:830–837.

15. Veselka J, Jensen M, Liebregts M et al. Alcohol septal ablation in patients with severe septal hypertrophy. Heart 2020;106:462–466.

16. Kimmelstiel C, Zisa DC, Kuttab JS et al. Guideline-Based Referral for Septal Reduction Therapy in Obstructive Hypertrophic Cardiomyopathy Is Associated With Excellent Clinical Outcomes. Circ Cardiovasc Interv 2019;12:e007673.

17. Veselka J, Faber L, Liebregts M et al. Short- and long-term outcomes of alcohol septal ablation for hypertrophic obstructive cardiomyopathy in patients with mild left ventricular hypertrophy: a propensity score matching analysis. Eur Heart J 2019;40:1681–1687.

18. Veselka J, Faber L, Jensen MK et al. Effect of Institutional Experience on Outcomes of Alcohol Septal Ablation for Hypertrophic Obstructive Cardiomyopathy. Can J Cardiol 2018;34:16–22.

19. Paul S. Alcohol septal ablation for obstructive hypertrophic cardiomyopathy: a word of balance. J Am Coll Cardiol 2017;70:489–494.

20. Liebregts M, Faber L, Jensen MK et al. Outcomes of Alcohol Septal Ablation in Younger Patients With Obstructive Hypertrophic Cardiomyopathy. JACC Cardiovasc Interv 2017;10:1134–1143.

21. Josef V, Morten Kvistholm J, Max L et al. Long-term clinical outcome after alcohol septal ablation for obstructive hypertrophic cardiomyopathy: results from the Euro-ASA registry. Eur Heart J 2016;37:1517–23.

22. Vriesendorp PA, Liebregts M, Steggerda RC et al. Long-term outcomes after medical and invasive treatment in patients with hypertrophic cardiomyopathy. JACC Heart Fail 2014;2:630–6.

23. Veselka J, Krejci J, Tomasov P, Zemanek D. Long-term survival after alcohol septal ablation for hypertrophic obstructive cardiomyopathy: a comparison with general population. Eur Heart J 2014;35:2040–5.

24. Sorajja P, Binder J, Nishimura RA et al. Predictors of an optimal clinical outcome with alcohol septal ablation for obstructive hypertrophic cardiomyopathy. Catheter Cardiovasc Interv 2013;81:E58–67.

25. Sorajja P, Ommen SR, Holmes DR, Jr. et al. Survival after alcohol septal ablation for obstructive hypertrophic cardiomyopathy. Circulation 2012;126:2374–80.

26. Michael AF, Ulrich S. Hypertrophic obstructive cardiomyopathy: alcohol septal ablation. Eur Heart J 2011;32:1059–64.

27. ten Cate FJ, Soliman OI, Michels M et al. Long-term outcome of alcohol septal ablation in patients with obstructive hypertrophic cardiomyopathy: a word of caution. Circ Heart Fail 2010;3:362–9.

28. Fernandes VL, Nielsen C, Nagueh SF et al. Follow-up of alcohol septal ablation for symptomatic hypertrophic obstructive cardiomyopathy the Baylor and Medical University of South Carolina experience 1996 to 2007. JACC Cardiovasc Interv 2008;1:561–70.

29. Kuhn H, Lawrenz T, Lieder F et al. Survival after transcoronary ablation of septal hypertrophy in hypertrophic obstructive cardiomyopathy (TASH): a 10 year experience. Clin Res Cardiol 2008;97:234–43.

30. Sorajja P, Valeti U, Nishimura RA et al. Outcome of alcohol septal ablation for obstructive hypertrophic cardiomyopathy. Circulation 2008;118:131–9.

31. Faber L, Welge D, Fassbender D, Schmidt HK, Horstkotte D, Seggewiss H. One-year follow-up of percutaneous septal ablation for symptomatic hypertrophic obstructive cardiomyopathy in 312 patients: predictors of hemodynamic and clinical response. Clin Res Cardiol 2007;96:864–73.

32. David RH, Uma SV, Rick AN. Alcohol septal ablation for hypertrophic cardiomyopathy: indications and technique. Catheter Cardiovasc Interv 2005;66:375–89.

33. Haran B, Ulrich S. Alcohol septal ablation for hypertrophic obstructive cardiomyopathy. 2005.

34. Bourque C, Reant P, Bernard A et al. Comparison of Surgical Ventricular Septal Reduction to Alcohol Septal Ablation Therapy in Patients with Hypertrophic Cardiomyopathy. Am J Cardiol 2022;172:109–114.

35. Nguyen A, Schaff HV, Hang D et al. Surgical myectomy versus alcohol septal ablation for obstructive hypertrophic cardiomyopathy: A propensity score-matched cohort. J Thorac Cardiovasc Surg 2019;157:306–315 e3.

36. Singh K, Qutub M, Carson K, Hibbert B, Glover C. A meta analysis of current status of alcohol septal ablation and surgical myectomy for obstructive hypertrophic cardiomyopathy. Catheter Cardiovasc Interv 2016;88:107–15.

37. Leonardi RA, Kransdorf EP, Simel DL, Wang A. Meta-analyses of septal reduction therapies for obstructive hypertrophic cardiomyopathy: comparative rates of overall mortality and sudden cardiac death after treatment. Circ Cardiovasc Interv 2010;3:97– 104.

38. Ralph-Edwards A, Woo A, McCrindle BW et al. Hypertrophic obstructive cardiomyopathy: comparison of outcomes after myectomy or alcohol ablation adjusted by propensity score. J Thorac Cardiovasc Surg 2005;129:351–8.

39. Olivotto I, Oreziak A, Barriales-Villa R et al. Mavacamten for treatment of symptomatic obstructive hypertrophic cardiomyopathy (EXPLORER-HCM): a randomised, double-blind, placebo-controlled, phase 3 trial. Lancet 2020;396:759–769.

40. Ho CY, Olivotto I, Jacoby D et al. Study Design and Rationale of EXPLORER-HCM: Evaluation of Mavacamten in Adults With Symptomatic Obstructive Hypertrophic Cardiomyopathy. Circ Heart Fail 2020;13:e006853.

41. Desai MY, Wolski K, Owens A et al. Mavacamten in Patients With Hypertrophic Cardiomyopathy Referred for Septal Reduction: Week 128 Results From VALOR-HCM. Circulation 2025;151:1378–1390.

42. Desai MY, Owens A, Wolski K et al. Mavacamten in Patients With Hypertrophic Cardiomyopathy Referred for Septal Reduction: Week 56 Results From the VALOR-HCM Randomized Clinical Trial. JAMA Cardiol 2023;8:968–977.

43. Desai MY, Owens A, Geske JB et al. Myosin Inhibition in Patients With Obstructive Hypertrophic Cardiomyopathy Referred for Septal Reduction Therapy. J Am Coll Cardiol 2022;80:95–108.

44. Garcia-Pavia P, Maron MS, Masri A et al. Aficamten or metoprolol monotherapy for obstructive hypertrophic cardiomyopathy. New England Journal of Medicine 2025;393:949–960.

45. Samhan A, Saleh D, Kim EY et al. Comparison of Alcohol Septal Ablation With Mavacamten in Obstructive Hypertrophic Cardiomyopathy. The American Journal of Cardiology 2025;239:51–56.

46. von Elm E, Altman DG, Egger M, Pocock SJ, Gøtzsche PC, Vandenbroucke JP. The Strengthening the Reporting of Observational Studies in Epidemiology (STROBE) statement: guidelines for reporting observational studies. The Lancet 2007;370:1453– 1457.

47. Sedaghat-Hamedani F, Kayvanpour E, Tugrul OF et al. Clinical outcomes associated with sarcomere mutations in hypertrophic cardiomyopathy: a meta-analysis on 7675 individuals. Clin Res Cardiol 2018;107:30–41.

48. Masri A, Lester SJ, Stendahl JC et al. Long-Term Safety and Efficacy of Mavacamten in Symptomatic Obstructive Hypertrophic Cardiomyopathy: Interim Results of the PIONEER-OLE Study. J Am Heart Assoc 2024;13:e030607.

49. Garcia-Pavia P, Oreziak A, Masri A et al. Long-term effect of mavacamten in obstructive hypertrophic cardiomyopathy. Eur Heart J 2024;45:5071–5083.

50. Maron MS, Masri A, Nassif ME et al. Aficamten for Symptomatic Obstructive Hypertrophic Cardiomyopathy. New England Journal of Medicine 2024;390:1849– 1861.

